# Proteomic Immune Signatures of Severe HIV-Associated Tuberculosis in Sub-Saharan Africa: A Prospective, Multicenter Analysis from Uganda

**DOI:** 10.64898/2025.12.31.25343299

**Authors:** Jesse E. Ross, Alin S. Tomoiaga, Nicholas Owor, Xuan Lu, Joseph Shinyale, Tonny Kiyingi, Ignatius Asasira, Peter James Eliku, John Bosco Nsubuga, Christopher Nsereko, Irene Nayiga, Stephen Kyebambe, Thomas Ochar, Moses Kiwubeyi, Rittah Nankwanga, Kai Nie, Hui Xie, Sam Miake-Lye, Bryan Villagomez, Jingjing Qi, Steven J Reynolds, Martina Cathy Nakibuuka, John Kayiwa, Mercy Haumba, Joweria Nakaseegu, Xiaoyu Che, Risa Hoffman, John A Belperio, Julius J. Lutwama, Seunghee Kim-Schulze, Max R. O’Donnell, Barnabas Bakamutumaho, Matthew J. Cummings

## Abstract

**Objective:** Severe tuberculosis (TB) is a major cause of critical illness and death in people living with HIV (PLWH) worldwide. Despite this, the immunopathology of severe HIV-associated TB (HIV/TB) is poorly understood. We aimed to identify an immunopathologic signature of severe HIV/TB in sub-Saharan Africa.

**Design and Setting:** We analyzed proteomic data from two prospective observational cohorts of adults hospitalized with severe undifferentiated infection in Uganda: an urban discovery cohort (Entebbe, N=241) and a rural validation cohort (Tororo, N=253).

**Patients:** Adults (age ≥18 years) hospitalized with severe febrile illness

**Interventions:** None

**Measurements and Main Results:** Across both cohorts, severe HIV-associated TB was common, affecting 18% of participants in the discovery cohort and 21% in the validation cohort. Overall mortality was significant (30-day mortality of 22% in the discovery cohort & 60-day mortality of 26% in the validation cohort). Participants were stratified into three HIV/TB phenotypes: HIV-negative without TB, PLWH without TB, and PLWH with microbiologically diagnosed TB. We applied ordinal random forest models in the discovery cohort to identify proteins strongly predictive of progressive HIV/TB phenotype. In both cohorts, PLWH with microbiologically diagnosed TB were at highest risk of critical illness and death (30-day mortality of 42% in the discovery cohort & 60-day mortality of 52% in the validation cohort). An eight-protein signature reliably distinguished this phenotype, reflecting mediators of macrophage/dendritic cell activation (LAMP3), NK- and T-cell stimulation and cytotoxicity (CD70, CRTAM), B-cell activation (IGLC2), protease-mediated tissue injury (PRSS2), dysregulated coagulation (SERPINA5), extracellular matrix remodeling (EFEMP1), and GH/IGF axis dysregulation (IGFBP3).

**Conclusions:** We identified an immunologic signature of severe HIV-associated TB defined by mediators of macrophage/dendritic cell and cytotoxic lymphocyte activation, extracellular matrix remodeling, and dysregulated coagulation. These findings offer new insight into HIV/TB pathobiology and highlight potential targets for host-directed therapies in this high-risk population.

**Key Points:** *Question:* What host-response patterns characterize severe HIV-associated tuberculosis among adults hospitalized with severe febrile illness in sub-Saharan Africa?

*Findings:* In two prospective cohorts of adults hospitalized with severe febrile illness in Uganda, severe HIV-associated tuberculosis accounted for 18-21% of cases and was associated with higher rates of physiological instability and mortality. An eight-protein host-response signature reproducibly distinguished this high-risk phenotype, reflecting immune activation, tissue injury, extracellular matrix remodeling, and dysregulated coagulation.

*Meaning:* Severe HIV-associated tuberculosis is associated with a distinct, high-risk clinical phenotype characterized by reproducible host-response patterns that may inform risk stratification and host-directed therapeutic strategies.

## Introduction

Severe tuberculosis (TB) is the leading cause of hospitalization and death among people living with HIV (PLWH) globally, with the highest TB burden in low- and middle-income countries (1–4). In sub-Saharan Africa (SSA), where HIV and TB are co-epidemic, MTB is the predominant cause of critical illness and death among hospitalized PLWH (5–6). In this population, the frequency of disseminated MTB infection can exceed 40%, with short-term mortality approaching 50% (6–9).

Despite this substantial burden, the immunopathology of severe HIV-associated tuberculosis (HIV/TB) remains poorly understood. Few studies have characterized multidimensional host responses in PLWH with severe TB, limiting insight into the mechanisms driving immune dysfunction in this critically ill population and, consequently, limiting the ability to target specific treatment mechanisms that may improve outcomes (1, 10–17). Available data suggests that MTB infection may be linked to activation of both pro- and anti-inflammatory innate immune pathways, dysregulated T-cell activation and exhaustion, endothelial dysfunction, and extracellular matrix remodeling (1, 10–17), features that partially overlap with those observed in both HIV and critically ill patients with non-TB bacterial infections (18–20). However, most studies to date have relied on narrow biomarker panels, single-center designs, and lacked comparator groups such as HIV-uninfected and TB-negative controls, which reduces immunologic resolution, hinders attribution of TB-specific responses versus HIV or other critical illness, and limits generalizability across settings (1, 10–17). Concurrently, emerging evidence from other severe infections underscores substantial heterogeneity in host immune responses, shaped not only by the pathogen but also by host-specific factors such as comorbidities and immune status (19–21). In PLWH, chronic immune activation, T-cell exhaustion, and endothelial dysfunction further influence host-pathogen interactions, potentially giving rise to distinct immunopathologic signatures of critical illness (23–28). This biological complexity underscores the need for higher-resolution immune profiling in severe HIV/TB.

To address these gaps, we conducted high-throughput serum proteomic profiling in two prospective hospital-based cohorts in Uganda to identify a reproducible immune signature of severe HIV-associated TB. We hypothesized that hospitalized PLWH with severe TB would exhibit a reproducible serum proteomic signature reflecting exaggerated innate immune activation and T-cell exhaustion.

## Methods

### Study Settings, Outcomes, and Participants

We analyzed data from two prospective observational cohorts of adults hospitalized with severe febrile illness at two public hospitals in Uganda (**Figure S1**). The discovery cohort (RESERVE-U-1-EBB) was enrolled at Entebbe Regional Referral Hospital (ERRH), an urban referral hospital in central Uganda, between April 2017 and August 2019. The validation cohort (RESERVE-U-2-TOR) was enrolled at Tororo General Hospital (TGH), a rural district hospital in eastern Uganda, between November 2021 and August 2023. Enrollment criteria were uniform across the RESERVE-U cohorts. Non-pregnant adults (age ≥18 years) hospitalized with severe febrile illness (i.e., severe undifferentiated infection) were enrolled based on a reported history of fever or axillary temperature ≥ 37.5°C and illness severe enough to warrant admission. The primary clinical outcomes for RESERVE-U-1-EBB and RESERVE-U-2-TOR were 30-and 60-day vital status, respectively. Details of these cohorts have been published and are included in the supplement along with further details of study sites, procedures, and outcomes (18).

### Study Procedures

Enrolled participants underwent standardized clinical assessments, including rapid testing for HIV by a Ministry of Health-recommended algorithm (29) and pathogens implicated as etiologies of severe infection in SSA, as informed by World Health Organization (WHO) guidelines (28). For PLWH, MTB testing included Xpert MTB/RIF Ultra (Cepheid, Sunnyvale, CA, USA) on sputum samples and Determine TB-LAM (Abbott/Alere, Lake Bluff, IL, USA) on urine samples, where specimens were obtainable. TB testing of patients without HIV was at the discretion of treating clinicians.

Participants from both cohorts were individually categorized into one of three ordinal groups reflecting HIV/TB co-infection phenotypes: (1) HIV-negative and TB-negative (HIV-/TB-), (2) HIV-positive and TB-negative (HIV+/TB-), (3) HIV-positive and TB-positive (HIV+/TB+). Microbiological diagnosis of TB was based on a positive sputum Xpert MTB/RIF Ultra or urine TB-LAM result. HIV-uninfected participants were classified at TB-negative if sputum Xpert MTB/RIF Ultra was negative and there was no clinical suspicion of TB. PLWH were classified as TB-negative if both sputum Xpert MTB/RIF Ultra and urine TB-LAM results were negative and there was no clinical suspicion of TB. In both cases, no clinical suspicion of TB was defined as the treating clinicians’ decision not to initiate empiric anti-TB therapy. Serum samples were collected at the time of study enrollment and stored at –80°C until the time of analysis.

### Olink Proteomics

Proteomic profiling was performed on cryopreserved serum samples using the Olink Target Immunooncology and Cardiometabolic panels (Olink Proteomics AB, Uppsala, Sweden), which together quantify concentrations of 184 soluble proteins (18). These panels were utilized to capture multiple biological domains implicated in the host response to severe infection (i.e., innate and adaptive immune activation, endothelial dysfunction, and dysregulated coagulation), including severe TB. For each panel, proteins were quantified via the Olink Proximity Extension Assay (PEA) platform, and relative protein abundance was expressed in log_2_ normalized protein expression (NPX) units. Proteins were excluded from downstream analyses if <20% of NPX values were above each panel’s estimated limit of detection (18).

### Biomarker Selection Using Ordinal Forest Models

To identify soluble proteins associated with HIV/TB phenotypes, we applied an ordinal random forest model to the RESERVE-U-1-EBB discovery cohort, using the three-level HIV/TB phenotype classification as the ordinal outcome. To mitigate class imbalance and optimize feature selection across all HIV/TB phenotypes, we performed class-balanced upsampling through duplicating existing minority class samples, thereby equalizing group representation during model training. Upsampling was restricted to the training data within each resampling split, and the independent validation set was never upsampled. Random forest model parameters included mtry = 5, nsets = 500, ntreeperdiv = 100, and nbest = 20. The predictive strength of each protein was quantified using permutation-based variable importance. The top ten proteins were selected from the importance rankings to provide a parsimonious set of features that balances model interpretability with sufficient depth for downstream analyses.

### Multivariable Analysis of Soluble Proteins and HIV/TB Phenotypes

To quantify associations between soluble proteins and HIV/TB phenotypes, we analyzed the top 10 proteins identified in the RESERVE-U-1-EBB discovery cohort using separate multivariable linear regression models. Each model included NPX-quantified protein expression as the continuous outcome and HIV/TB phenotype as an ordinal predictor. Models were adjusted for age, sex, illness duration prior to enrollment, and malaria status. To avoid collider bias, we did not adjust for physiological severity, which may be influenced by both HIV/TB infection and host responses. We reported regression coefficients, 95% confidence intervals, and p-values for linear trend, correcting the latter for multiple testing using the Benjamini-Hochberg false discovery rate (FDR) method. Proteins with FDR-adjusted p-values <0.05 in the RESERVE-U-1-EBB discovery cohort were carried forward for testing in the RESERVE-U-2-TOR validation cohort using identical regression models.

To explore whether proteomic associations with HIV/TB phenotypes were independent of HIV-related immunosuppression and/or viremia, we conducted a sensitivity analysis restricted to PLWH in the RESERVE-U-2-TOR cohort. We fit multivariable linear regression models for each of the top ten proteins, with NPX-quantified protein expression as the continuous outcome and HIV/TB phenotype as the ordinal predictor, adjusted for CD4 count and HIV-1 viral load. CD4 count was modeled as a continuous variable while HIV-1 viral load was modeled as a binary indicator: suppressed versus unsuppressed. Details of CD4 count and HIV-1 viral load quantification methods have been described and are in the supplement (**Table S1**) (18). Age, sex, illness duration, and malaria status were not included in this model to prevent overfitting and underpowering of this HIV-restricted exploratory analysis. Similarly, given the smaller sample size and exploratory intent, FDR correction was not applied and unadjusted p-values are reported. CD4 count and HIV-1 viral load data were unavailable from RESERVE-U-1-EBB participants.

All analyses were conducted in R version 4.3.2 using the tidyverse, ordinalForest, caret, stats, broom, and patchwork packages.

### Ethics Statement

Each enrolled participant or their surrogate provided written informed consent. Study protocols were approved by ethics committees at Columbia University (AAAR1450), Uganda Virus Research Institute (GC/127/17/02-06/582), and Uganda National Council for Science and Technology (HS2308).

## Results

### Participant Characteristics

A total of 494 participants were analyzed: 241 in the RESERVE-U-1-EBB (discovery) cohort and 253 in the RESERVE-U-2-TOR (validation) cohort. Characteristics of analyzed patients stratified by HIV/TB phenotype are presented in **Tables 1 and 2**. Approximately half of participants were female. The median age was 32 years (IQR 26 – 40) in Entebbe and 48 years (IQR 33 – 61) in Tororo. The proportion of PLWH was similar in Entebbe and Tororo (51% versus 50% respectively), though the proportion of PLWH receiving anti-retroviral therapy was higher in Tororo (83% versus 68%). In PLWH enrolled in RESERVE-U-2-TOR, the median CD4 count was 350 cells/µL (IQR 300-600) and 71% had suppressed HIV-1 viral loads (<150 copies/mL). The frequency of microbiologically diagnosed TB was similar across sites: 21% in Tororo and 18% in Entebbe. Among those with microbiologically diagnosed TB, disseminated disease as indicated by positive urine TB-LAM testing was present in 40/47 (85%) of PLWH in Entebbe and 51/53 (96%) of PLWH in Tororo. Quick Sequential Organ Failure Assessment (qSOFA) score greater than or equal to 1, previously shown to predict infection-related mortality in low- and middle-income countries, was present in 87% and 88% of participants in the discovery and validation cohorts, respectively (30–32). Mortality at 30- and 60-days was 22% in RESERVE-U-1-EBB and 26% in RESERVE-U-2-TOR, respectively. Most deaths occurred after hospital discharge. In both cohorts, PLWH with microbiologically diagnosed TB were at highest risk of physiological instability and death.

**Table 1:** Participant characteristics stratified by HIV/TB phenotype in RESERVE-U-1-EBB cohort.

| **Characteristics<sup>a</sup>** | **All participants**<br>**N = 241** | **HIV - / TB -**<br>**N = 117** | **HIV + / TB -**<br>**N = 77** | **HIV + / TB +**<br>**N=47** |
| --- | --- | --- | --- | --- |
| **Demographics** |  |  |  |  |
| Female sex | 149/241 (62%) | 71/117 (61%) | 52/77 (68%) | 26/47 (55%) |
| Age, years | 32 (26, 40) | 28 (23, 38) | 37 (30, 43) | 32 (27, 40) |
| Mid-upper arm circumference, cm | -- | -- | -- | -- |
| Illness duration prior to enrollment, days | 4 (2, 6) | 3 (2, 5) | 4 (3, 7) | 6 (3, 7) |
| **Clinical signs and physiological severity scores** |  |  |  |  |
| KPS at hospital admission | -- | -- | -- | -- |
| Whole-blood lactate, mmol/L | -- | -- | -- | -- |
| qSOFA score $\geq 1^b$ | 209/241 (87%) | 97/117 (83%) | 67/77 (87%) | 45/47 (96%) |
| qSOFA score $\geq 2^b$ | 102/241 (42%) | 30/117 (26%) | 39/77 (51%) | 33/47 (70%) |
| Modified Early Warning Score | 3 (2, 4) | 3 (2, 3) | 3 (2, 4) | 5 (3, 6) |
| Universal Vital Assessment score | 2 (1, 4) | 1 (0, 2) | 3 (2, 5) | 3 (3, 7) |
| **HIV Characteristics** |  |  |  |  |
| PLWH | 124/241 (51%) | NA | 77/77 (100%) | 47/47 (100%) |
| WHO HIV clinical stage 3 or 4 | 97/124 (78%) | NA | 50/77 (65%) | 47/47 (100%) |
| Receiving ART before hospitalization | 73/124 (59%) | NA | 45/77 (58%) | 28/47 (60%) |
| CD4 cell count (cells/mm<sup>3</sup>) | -- | -- | -- | -- |
| HIV-1 viral suppression | -- | -- | -- | -- |
| **Pathogen diagnostics** |  |  |  |  |
| Malaria RDT positive | 54/237 (23%) | 30/116 (26%) | 20/76 (26%) | 4/45 (8.9%) |
| Urine TB-LAM positive if PLWH | 40/99 (40%) | NA | NA | 40/47 (85%) |
| Cryptococcal ag. positive if PLWH | 9/29 (31%) | NA | 6/13 (46%) | 3/16 (19%) |
| Influenza PCR positive | 14/219 (6.4%) | 8/106 (7.5%) | 5/71 (7.0%) | 1/42 (2.4%) |
| **Outcomes** |  |  |  |  |
| Death in hospital | 14/241 (5.8%) | 4/117 (3.4%) | 4/77 (5.2%) | 6/47 (13%) |
| Death at 30 days | 49/219 (22%) | 10/106 (9.4%) | 20/68 (29%) | 19/45 (42%) |

**Abbreviations:**, KPS: Karnofsky Performance Status, qSOFA: quick Sepsis-related Organ Failure Assessment, PLWH: person living with HIV; WHO: World Health Organization, ART: anti-retroviral therapy, RDT: rapid diagnostic test, TB: tuberculosis, LAM: lipoarabinomannan, PCR: polymerase chain reaction
**Legend:** <sup>a</sup>Dichotomous and categorical variables summarized as counts (proportions), continuous variables summarized as medians (interquartile ranges), n if fewer patients with available data than the total number of patients in the cohort; <sup>b</sup>Systolic blood pressure $\leq 100$ mmHg, respiratory rate $\geq 22$ breaths/min, and altered mental status, latter defined using

**Table 2:** Participant characteristics stratified by HIV/TB phenotype in RESERVE-U-2-TOR cohort.

| **Characteristics<sup>a</sup>** | **All participants**<br>**N = 253** | **HIV - / TB -**<br>**N = 126** | **HIV + / TB -**<br>**N = 73** | **HIV + / TB +**<br>**N=54** |
| --- | --- | --- | --- | --- |
| **Demographics** |  |  |  |  |
| Female sex | 158/253 (62%) | 84/126 (67%) | 39/73 (53%) | 35/54 (65.0%) |
| Age, years | 48 (33, 61) | 48 (25, 70) | 47 (40, 56) | 48 (40, 56) |
| Mid-upper arm circumference, cm | 24 (22, 26) | 24 (23, 26) | 24 (23, 26) | 22 (20, 24) |
| Illness duration prior to enrollment, days | 6 (4, 10) | 6 (4, 9) | 7 (4, 10) | 9 (4, 14) |
| **Clinical signs, lactate, and physiological severity scores** |  |  |  |  |
| KPS at hospital admission | 60 (20, 70) | 70 (20, 80) | 60 (40, 70) | 50 (20, 70) |
| Whole-blood lactate, mmol/L | 2.4 (1.8, 3.0) | 2.4 (1.8, 3.0) | 2.4 (1.7, 2.9) | 2.4 (2.0, 3.0) |
| qSOFA score $\geq 1^b$ | 222/253 (88%) | 110/126 (87%) | 65/73 (89%) | 47/54 (87%) |
| qSOFA score $\geq 2^b$ | 100/253 (40%) | 46/126 (37%) | 29/73 (40%) | 25/54 (46%) |
| Modified Early Warning Score | 3 (2, 5) | 3 (2, 5) | 4 (2, 5) | 3 (2, 5) |
| Universal Vital Assessment score | 2 (1, 5) | 1 (0, 4) | 3 (2, 6) | 4 (2, 6) |
| **HIV Characteristics** |  |  |  |  |
| PLWH | 127/253 (50%) | NA | 73/73 (100%) | 54/54 (100%) |
| WHO HIV clinical stage 3 or 4 | 93/127 (73%) | NA | 51/73 (70%) | 42/54 (78%) |
| Receiving ART before hospitalization | 105/127 (83%) | NA | 57/73 (78%) | 48/54 (89%) |
| CD4 count, cells/mm<sup>3</sup> | 350 (300, 600) | NA | 400 (300, 600) | 344 (200, 500) |
| HIV-1 viral suppression | 85/120 (71%) | NA | 49/68 (72%) | 36/52 (69%) |
| **Pathogen diagnostics** |  |  |  |  |
| Malaria RDT positive | 74/253 (29%) | 41/126 (33%) | 18/73 (25%) | 15/54 (28%) |
| Urine TB-LAM positive | 51/122 (42%) | NA | 0/69 (0%) | 51/53 (96%) |
| Cryptococcal ag. positive if PLWH | 10/123 (8.1%) | NA | 6/70 (8.6%) | 4/53 (7.5%) |
| Influenza PCR positive | 14/246 (5.7%) | 9/123 (7.3%) | 5/72 (6.9%) | 0/51 (0%) |
| SARS-CoV-2 PCR positive | 13/246 (5.3%) | 8/123 (6.5%) | 1/72 (1.4%) | 4/51 (7.8%) |
| **Outcomes** |  |  |  |  |
| Death in hospital | 5/253 (2.0%) | 1/126 (0.8%) | 1/73 (1.4%) | 3/54 (5.6%) |
| Death at 30 days | 51/252 (20%) | 11/126 (8.7%) | 15/72 (21%) | 25/54 (46%) |
| Death at 60 days | 65/252 (26%) | 18/126 (14%) | 19/72 (26%) | 28/54 (52%) |

Abbreviations., KPS: Karnofsky Performance Status, qSOFA: quick Sepsis-related Organ Failure Assessment, PLWH: person living with HIV; WHO: World Health Organization, ART: anti-retroviral therapy, RDT: rapid diagnostic test, TB: tuberculosis, LAM: lipoarabinomannan, PCR: polymerase chain reaction
Legend: <sup>a</sup>Dichotomous and categorical variables summarized as counts (proportions), continuous variables summarized as medians (interquartile ranges), n if fewer patients with available data than the total number of patients in the cohort; <sup>b</sup>Systolic blood pressure $\leq 100$ mmHg, respiratory rate $\geq 22$ breaths/min, and altered mental status, latter defined using

### Biomarker Results

After excluding 11 proteins due to high proportions of expression values below each Olink panel’s estimated limits of detection, we applied an ordinal random forest model in the RESERVE-U-1-EBB discovery cohort to 173 soluble proteins. The top ten most important proteins for prediction of HIV/TB phenotype reflected macrophage and dendritic cell activation (LAMP3), NK and T-cell stimulation and cytotoxic function (CD70, CRTAM, IL-18), B cell activation and immunoglobulin production (IGLC2), protease-mediated tissue injury (PRSS2), dysregulated coagulation (SERPINA5), leukocyte adhesion and immune cell trafficking (ICAM3), extracellular matrix remodeling (EFEMP1), and growth hormone (GH) / insulin-like growth factor (IGF) axis dysregulation (IGFBP-3) (**Figure S2**).

In the RESERVE-U-1-EBB discovery cohort, all ten proteins were significantly associated with HIV/TB co-infection phenotype in multivariable linear regression models, including after FDR adjustment (**Figure 1A, Table S2**). Expression levels of eight proteins increased across the HIV-/TB-, HIV+/TB-, and HIV+/TB+ groups, while SERPINA5 and IGFBP-3 demonstrated an inverse association, with lower expression in participants with severe HIV/TB. In the validation cohort (RESERVE-U-2-TOR), eight of the ten proteins remained significantly associated with HIV/TB co-infection phenotype (**Figure 1B, Table S3**); associations for IL-18 and ICAM3 did not reach statistical significance. Visual inspection of boxplots confirmed consistent expression gradients across the HIV-/TB-, HIV+/TB-, and HIV+/TB+ groups in both cohorts (**Figures 2A and 2B**).

**Figure 1A.**
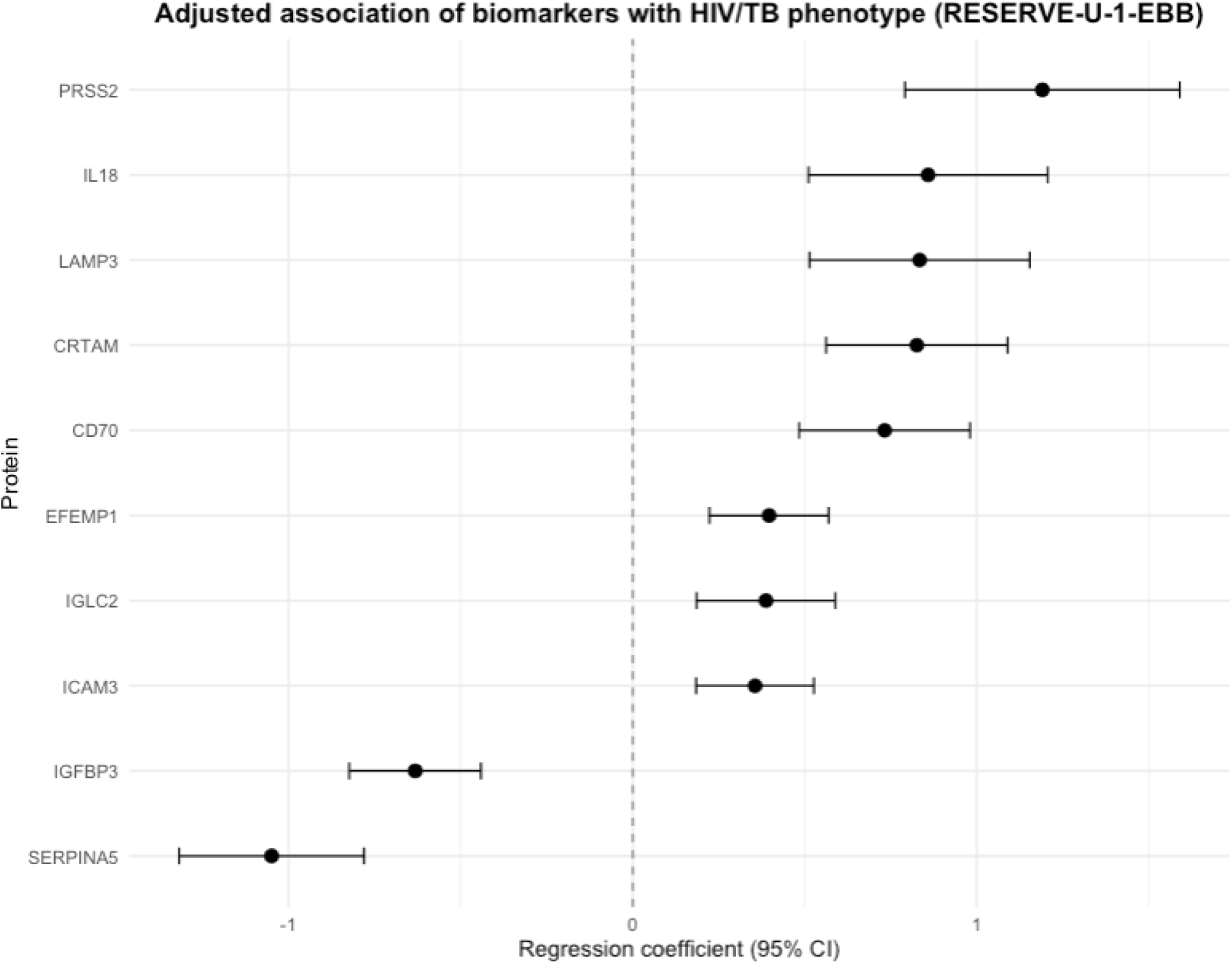
Forest plot showing adjusted regression coefficients and 95% confidence intervals (CI) for the association between HIV/TB phenotype and protein expression in the RESERVE-U-1-EBB discovery cohort. Coefficients and 95% CIs generated from linear regression models including NPX-quantified protein expression as the continuous outcome and HIV/TB phenotype as an ordinal predictor, adjusted for age, sex, illness duration prior to enrollment, and malaria status (N = 241).

**Figure 1B.**
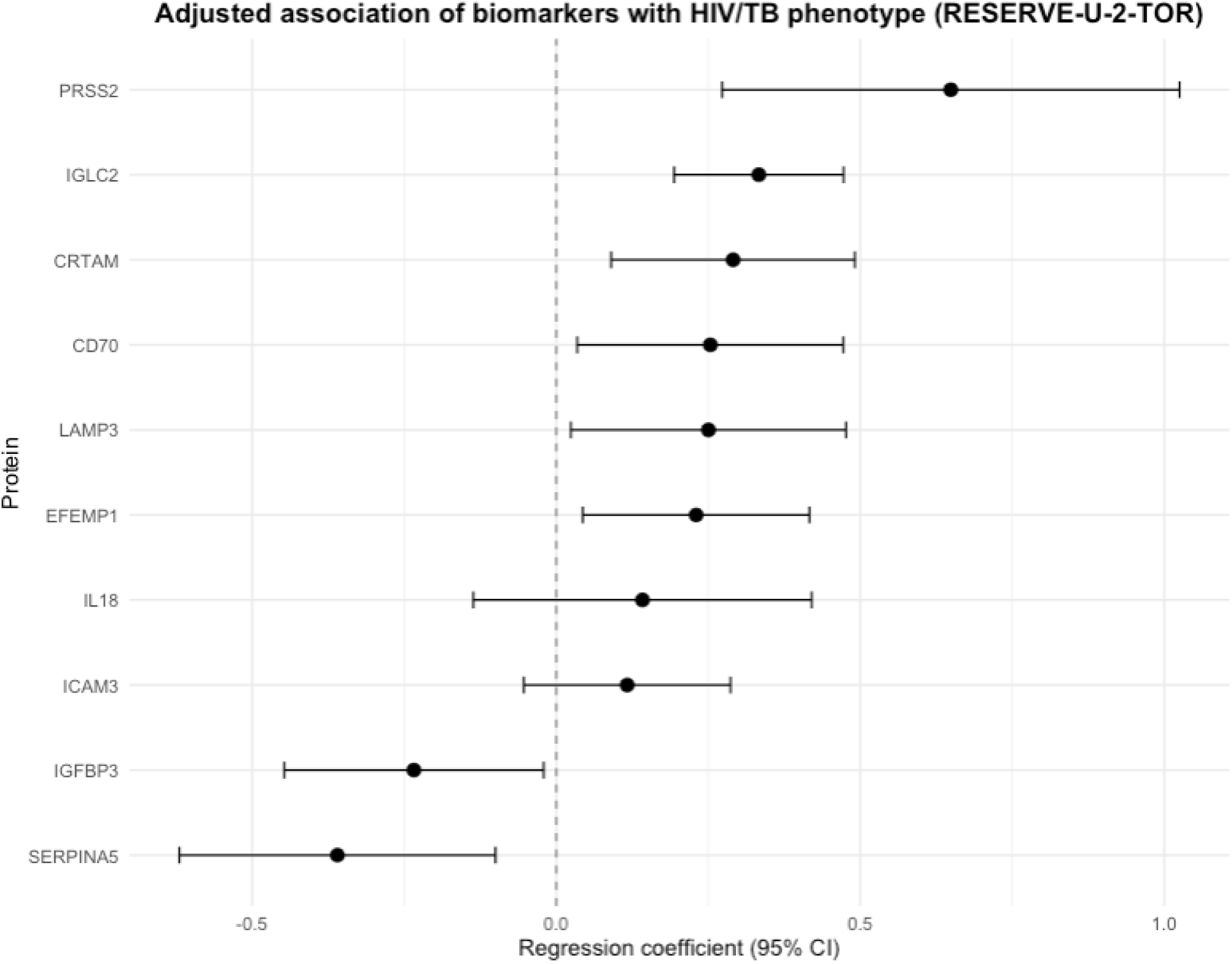
Forest plot showing adjusted regression coefficients and 95% confidence intervals (CI) for the association between progressive HIV/TB phenotype and protein expression in the RESERVE-U-2-TOR validation cohort. Coefficients and 95% CIs generated from linear regression models including NPX-quantified protein expression as the continuous outcome and HIV/TB phenotype as an ordinal predictor, adjusted for age, sex, illness duration prior to enrollment, and malaria status (N = 253).

**Figure 2A.**
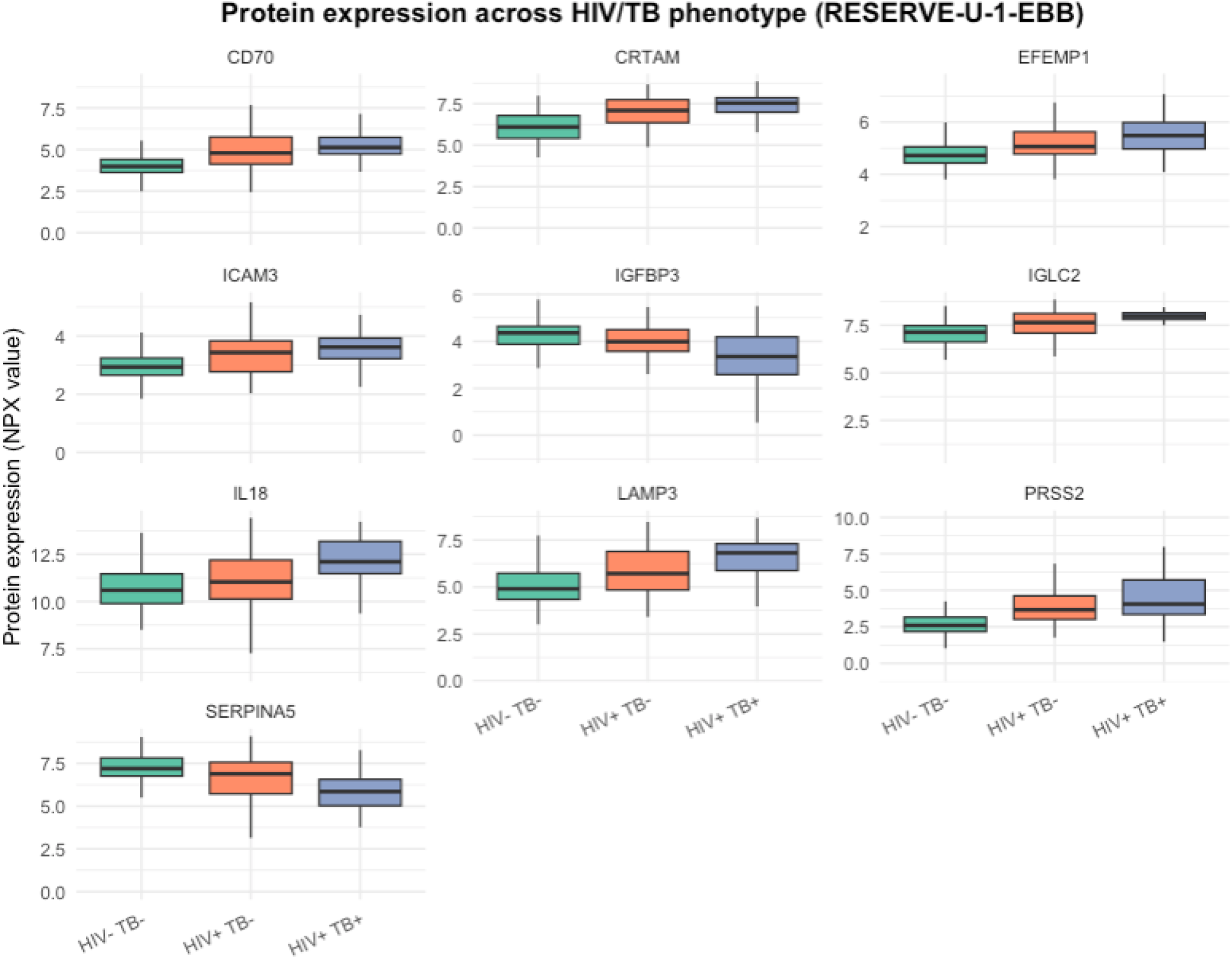
Boxplots showing distribution of top 10 most predictive proteins across HIV/TB phenotypes in RESERVE-U-1-EBB discovery cohort (N = 241).

**Figure 2B.**
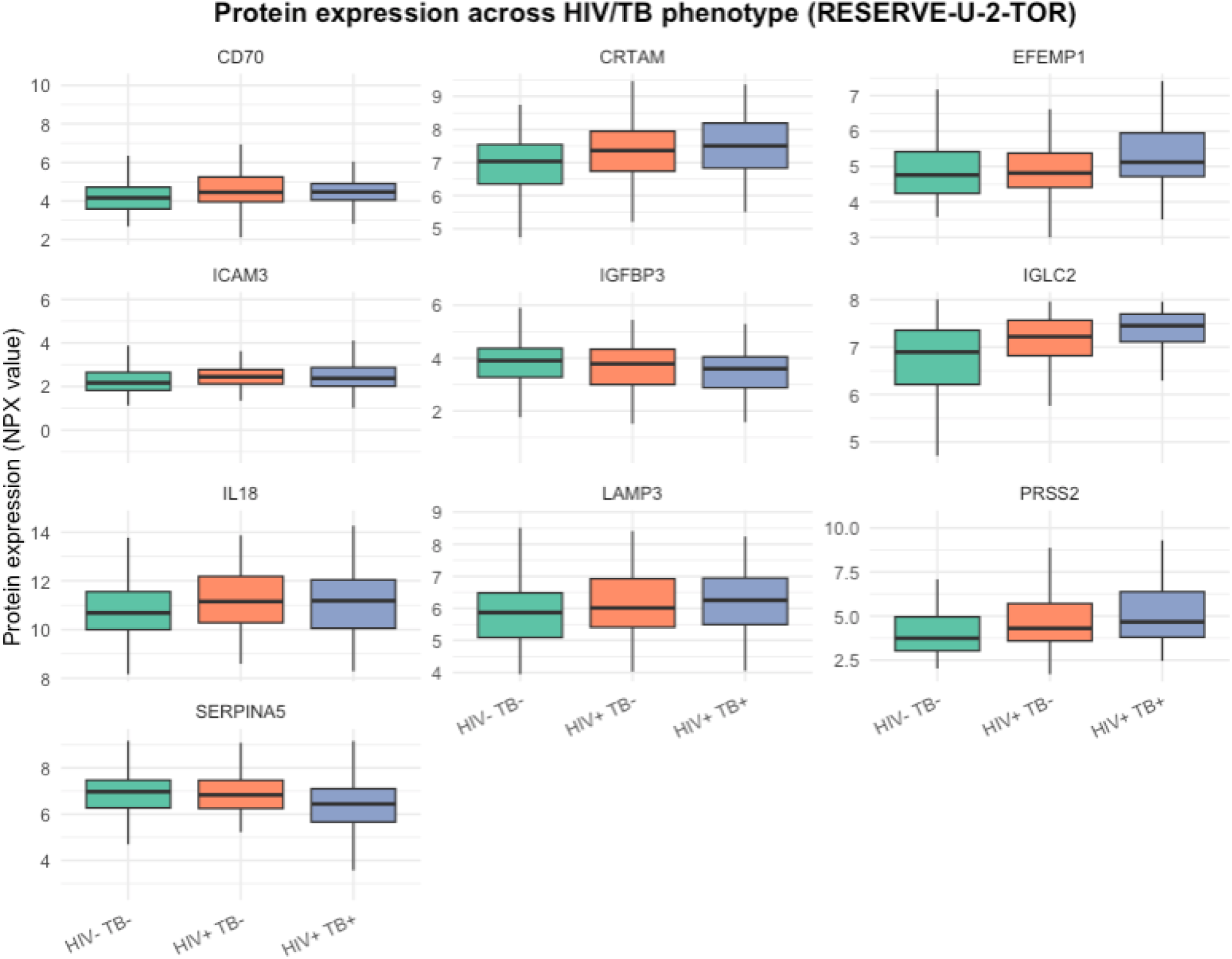
Boxplots showing distribution of top 10 most predictive proteins across HIV/TB phenotypes in RESERVE-U-2-TOR validation cohort (N = 253).

In exploratory linear regression models restricted to PLWH in RESERVE-U-2-TOR and adjusted for CD4 count and HIV-1 viral load (N = 127), three proteins, EFEMP1, IGLC2, and SERPINA5, remained significantly associated with HIV/TB co-infection phenotype (**Figure S3, Table S4**).

## Discussion

We identified and validated a proteomic immune signature associated with clinical phenotypes of HIV/TB in critically ill hospitalized adults in Uganda. To our knowledge, this is the first multicenter study in SSA to apply high-throughput proteomics to define and validate an immune signature of severe HIV-associated TB. These results advance translational understanding of immune dysregulation in a leading cause of critical illness and death among PLWH in the region. They also provide a foundation for biologically-driven stratification of severe HIV-associated TB and could inform the design of biomarker-guided treatment strategies for this high-risk population.

Our findings align with contemporary models of TB pathogenesis that emphasize the balance between protective and pathological inflammation (33, 34). In the context of HIV, loss and dysfunction of CD4 T cells and antigen-presenting cells (APCs) compromise immune containment, permitting bacillary dissemination and multiorgan injury (33, 34). The immune signature identified in our study supports this framework with *in vivo* data from the global HIV/TB epicenter: LAMP3 reflects maturation/activation of macrophages and dendritic cells and heightened antigen presentation; CD70 and CRTAM reflect cytotoxic lymphocyte stimulation and potential exhaustion; IGLC2 denotes B-cell activation and heightened humoral response; PRSS2 and EFEMP1 implicate proteolytic tissue injury and extracellular matrix remodeling; lower SERPINA5 is consistent with unrestrained serine-protease activity and coagulation dysregulation leading to an immune-thrombotic milieu; and lower IGFBP-3 is associated with GH/IGF axis dysregulation, a pattern previously reported with AIDS-associated wasting and increasing severity of sepsis (35, 36). Collectively, these features define a reproducible HIV/TB signature linking APC activation, cytotoxic and humoral lymphocyte stimulation, tissue injury, disordered coagulation, and GH/IGF-axis perturbation.

Prior studies from South Africa have highlighted the complexity of immune dysregulation in PLWH with severe TB, characterized by simultaneous activation and exhaustion across innate and adaptive pathways (1, 11, 15). Using relatively low-dimensional cytokine profiling, one study showed that non-survivors exhibited elevated concentrations of both pro-inflammatory (CSF-3, TNF, IL-6) and anti-inflammatory (IL-1Ra) cytokines, alongside expansion of pro-inflammatory monocytes but impaired capacity of innate cells to mount cytokine responses to bacterial antigens (15). Broader cytokine panels have identified a clinically high-risk profile dominated by innate and chemotactic mediators (IL-6, IL-1Ra), whereas T-cell–associated mediators were inversely correlated with mortality (1). More recently, anemia, upregulation of matrix metalloproteinases (MMPs), and extracellular matrix turnover have also been linked with disease severity and poor outcomes in severe HIV/TB (11, 13).

A complementary study from South Africa in PLWH quantified circulating MTB burden using blood-based Xpert MTB/RIF Ultra and demonstrated a dose–response relationship between bacillary load, host-response signatures of inflammation and tissue damage, and mortality risk (12). Related work from our group in Uganda showed that higher urine TB-LAM grade, a marker of bacillary burden, was significantly associated with increased pro-inflammatory innate and T-cell activation and chemotaxis (10). Building on these findings, our 173-protein analysis provides a higher-resolution view of HIV/TB pathogenesis, revealing axes of immune dysregulation not captured by narrower cytokine panels. Additionally, our work demonstrates external validation across two hospitals with HIV-negative and TB-negative comparators, improving dissection of HIV/TB-specific biology across the clinical spectrum. Together, these findings point to a coordinated and multifaceted host response involving both innate and adaptive pathways, building on prior studies implicating immune activation in TB severity and poor outcomes among PLWH. In conjunction with pathogen-centric work, this motivates a combined pathogen-load plus host-response staging framework to inform risk stratification and potentially therapeutic trial design in severe HIV/TB.

Across the clinical spectrum of HIV/TB co-infection, the respective contributions of HIV and MTB to immune dysregulation, including mechanisms that drive illness severity, remain poorly defined (34, 37). Prior studies from South Africa have shown that HIV viremia and CD4 lymphocytopenia are associated with host inflammatory responses, with IL-17A, which was not included in our Olink panels, emerging as a central immunological mediator (33, 34, 37). In contrast, a prior study from Malawi showed that blunted TNF production in monocytes, suggesting innate immunoparalysis, was strongly predictive of poor outcomes in severe TB while HIV co-infection was not (17). In an exploratory analysis restricted to PLWH in the RESERVE-U-2-TOR cohort, three proteins including IGLC2, SERPINA5, and EFEMP1 remained significantly associated with HIV/TB phenotype after adjustment for CD4 count and HIV-1 viral load. These persistent associations suggest that although HIV-related immunosuppression and/or viremia may contribute to immune dysregulation observed in our cohorts, it does not fully account for the proteomic signature observed in severe HIV/TB. Nonetheless, given the exploratory nature of this analysis, our results should be interpreted cautiously and considered hypothesis-generating.

Beyond identifying an immune signature of severe HIV/TB, our findings advance understanding of TB-specific immune responses, in contrast to those observed in other severe HIV-associated infections (18–20). Prior analyses of the RESERVE-U cohorts identified two pathobiologically-driven “Uganda Sepsis Signatures” (USS-1/USS-2), with USS-2, enriched for severe HIV/TB and higher mortality, defined by myeloid cell and inflammasome activation, T-cell exhaustion, endothelial barrier dysfunction and prothrombotic imbalance (18). Our findings build on these insights, suggesting the presence of a reproducible immunopathological signature specific to severe HIV-associated TB in the global sepsis epicenter.

Our findings have direct relevance for the development of host-directed therapeutics in severe HIV/TB. As diagnostic capabilities expand, protein-based signatures such as ours could enable precise, biomarker-guided treatment strategies and support the design of immunomodulatory treatment trials (12, 38). Several candidate therapies are already being evaluated, including in randomized controlled trials. For example, corticosteroids in NewStrat-TB aim to attenuate hyperinflammatory immune activation; doxycycline, a known MMP inhibitor, may mitigate matrix injury and remodeling, as suggested by preclinical and phase 2 data; aspirin targets the immunothrombotic axis, with signals of safety and potential benefit demonstrated in TB meningitis (11, 39–42). Conceptually, a parsimonious protein signature could be operationalized to enrich randomization in ongoing or future host-directed treatment trials and serve as an early biomarker endpoint. Moving forward, combining longitudinal proteomic profiling with quantitative pathogen measurements could jointly stage bacillary burden and host response. In a serial sampling study of disseminated HIV-associated TB, MTB DNA remained detectable in blood and urine for up to 14 days after therapy initiation (43), underscoring the sustained disseminated burden early in treatment and the potential value of pairing dynamic pathogen and host-response data to refine therapeutic strategies.

Our study has several strengths. Our primary analysis was performed in two independent prospective cohorts, across distinct geographic and temporal settings, encompassing nearly 500 hospitalized adults. These cohorts reflect real-world clinical populations at high risk for HIV/TB co-infection and mortality, enhancing the translational relevance of our findings. We employed a tiered analytical approach combining robust machine learning–based feature selection with prespecified multivariable models and validation. By treating HIV/TB phenotype as an ordinal variable, our models captured gradations in clinical and microbiological phenotypes, rather than relying on binary group comparisons. Predictive associations largely held across cohorts, increasing generalizability in high-burden settings. The Olink platform enabled relatively high-throughput measurement of soluble proteins across multiple biological domains. Importantly, several proteins retained significant associations even after adjustment for CD4 count and viral load, suggesting components of the signature reflect TB-specific immunopathology. Together, these strengths support the biological plausibility, scalability, and external validity of the identified immune signature.

Our study also has limitations. Although the two cohorts were geographically and temporally distinct, we did not conduct external validation outside Uganda. However, participant characteristics were broadly comparable to those of comparable cohorts in SSA (44). While we included only microbiologically confirmed TB cases in the HIV+/TB+ group, undiagnosed or subclinical TB may have been present in the HIV-/TB-and HIV+/TB- groups, potentially resulting in misclassification and attenuation of observed differences biasing results towards the null. Although we attempted to adjust for co-infections where data were available, including malaria, adjustment for cryptococcal and other bacterial infections was not possible due to limited testing. Blood cultures were not performed because of resource limitations. Therefore, residual confounding from unmeasured or incompletely characterized infections is possible, and our signature may reflect a more complex infectious background not limited to TB and HIV. We also did not include a direct measure of circulating mycobacterial burden, which could have strengthened our interpretation of immune activation patterns. Additionally, CD4 count and HIV-1 viral load data were only available in the RESERVE-U-2-TOR cohort, limiting adjustment for HIV disease severity in the discovery cohort. CD4 counts were multiply imputed in a proportion of PLWH in RESERVE-U-2-TOR (18). However, we believe this approach is justified, as simulation studies have demonstrated that multiple imputation reduces bias compared to complete case analysis, even at higher levels of missingness than our CD4 data (45). Proteomic data were collected at a single time point, which limits our ability to assess dynamic immune responses over the course of illness or treatment. Finally, our analysis focused on soluble protein expression, which may not fully reflect tissue-level immune processes, particularly in lung or intra-cellular compartments central to TB pathogenesis. Moreover, the Olink platform provides relative (NPX) assessments of protein expression rather than absolute quantification, which may limit cross-platform comparability.

## Conclusions

In conclusion, we identified a reproducible eight-protein immune signature associated with HIV/TB co-infection among critically ill hospitalized adults in Uganda. These findings provide new insights into the immunopathology of severe HIV/TB in this high-risk population and highlight potential candidate biomarkers that could support future prognostic and therapeutic research.

## Data Availability

Data sharing
Proteomic data are available in Dryad at https://doi.org/10.5061/dryad.b2rbnzsq2. Analytic code will be deposited in Github prior to publication.

https://doi.org/10.5061/dryad.b2rbnzsq2.

## Author Contributions

JER, BB, MRO’D, and MJC conceived the study and design. JJL, NO, CN, IN, SK, TO, MK, RN, JK, MH, and JN collected, organized and entered clinical data and blood samples and contributed to pathogen diagnostics. SKS, KN, HX, SML, BV contributed to proteomic analyses. JER, MJC, AT, and XL had full access to all data in the study and contributed to data cleaning, processing, verification, and analysis. JER developed the first draft of the manuscript. JER, AT, NO, XL, JS, TK, IA, PJE, JBN, CN, IN, SK, TO, MK, RN, KN, HX, SML, BV, JQ, SJR, MCN, JK, MH, JN, XC, WIL, RH, JAP, JJL, SKS, MRO, BB, MJC reviewed and edited the final version of the manuscript. JER had final responsibility for the decision to submit for publication.

## Declaration of Interests

MJC reports consulting fees from Vertex Pharmaceuticals and Veracyte unrelated to the submitted work. The remaining authors declare no conflicts of interest.

## Data sharing

Proteomic data are available in Dryad at https://doi.org/10.5061/dryad.b2rbnzsq2. Analytic code will be deposited in Github prior to publication.

## Funding

This work was supported by the National Heart Lung and Blood Institute (5T32HL105323-08 to J.E.R), the National Institute of Allergy and Infectious Diseases (K23AI16334 and R01AI184997 to M.J.C and the Division of Intramural Research [S.J.R]), and the National Center for Advancing Translational Sciences (UL1TR001873 to Columbia University, sub-award to M.R.O), National Institutes of Health. Additional support was provided by the Burroughs Wellcome Fund /American Society of Tropical Medicine and Hygiene (M.J.C), the Potts Memorial Foundation (J.E.R), and the Stony Wold-Herbert Fund, Inc. (J.E.R). This research was support in part by the Intramural Research Program of the NIH. The contributions of the NIH author(s) were made as part of their official duties as NIH federal employees, are in compliance with agency policy requirements, and are considered Works of the United States Government. However, the findings and conclusions presented in this paper are those of the author(s) and do not necessarily reflect the views of the NIH or the U.S. Department of Health and Human Services.

## Notes

### Competing Interest Statement

The authors have declared no competing interest.

### Summary of Updates

I have updated the abstract according to a specific journals guidelines, including a key points section. I have added the following sentence "Quick Sequential Organ Failure Assessment (qSOFA) score greater than or equal to 1, previously shown to predict infection-related mortality in low- and middle-income countries, was present in 87% and 88% of participants in the discovery and validation cohorts, respectively" I have added qSOFA greater than or equal to 1 in Tables 1 and 2. I have also adapted figures and supplemental material for journal guidelines.

