## Supplement for "Proteomic Immune Signatures of Severe HIV-Associated Tuberculosis in Sub-Saharan Africa: A Prospective, Multicenter Analysis from Uganda"

**Supplemental Methods**

Study Sites and Capacity

Entebbe Regional Referral Hospital (ERRH) is a 200-bed public regional referral hospital located in a peri-urban area of central Uganda. ERRH has a catchment area of approximately 4 million people. HIV prevalence in the catchment area is approximately 6% and malaria is mesoendemic. There is no high-dependency or intensive care unit at ERRH, and no vasopressor or inotropic agents are available on the hospital wards. Clinical hematology and chemistry testing is intermittently available in the hospital laboratory. During the RESERVE-U-1-EBB study period (April 2017-August 2019), no pressurized oxygen was available at ERRH, and oxygen concentrators were provided to hospital wards as part of the study program.

Tororo General Hospital (TGH) is a 200-bed public district hospital located in a rural area of eastern Uganda. TGH has a catchment area of approximately 500,000 people. HIV prevalence in the catchment area is approximately 4% and malaria is hyperendemic. There is no high-dependency or intensive care unit at TGH, and no vasopressor or inotropic agents are available on the hospital wards. Clinical hematology and chemistry testing is intermittently available in the hospital laboratory. During the RESERVE-U-2-TOR study period (November 2021-August 2023) oxygen therapy was typically provided via nasal cannula and simple or non-rebreathing facemask using concentrators and/or cylinders. Concentrators were provided to hospital wards as part of the study program and cylinders were transported to TGH from an oxygen plant at the closest regional referral hospital.

Enrollment Criteria

Patients were included in the RESERVE-U studies if they fulfilled the following criteria: (1) age ≥18 years, (2) reported a history of fever or had a recorded axillary temperature of ≥37.5ºC at presentation, (3) had clinical illness severe enough to warrant admission to hospital (as per treating clinicians), and (4) were able to provide informed consent or had a surrogate available to do so. Patients were excluded if they presented following trauma, were known to be pregnant, or were admitted to a non-medical ward. During the study periods, consecutive admissions to the medical wards were screened for eligibility by study staff. Patients were screened on weekdays during daytime hours and were enrolled as close to admission as possible. Although patients 5-17 years of age were also included in the parent RESERVE-U cohorts, given variations in HIV and tuberculosis (TB)-specific infection risk and infection-related host responses across age groups, for this study's analyses we included only adults (age ≥18 years).

<u>Outcomes</u>

The primary clinical outcome of the RESERVE-U-1-EBB study was vital status at 30 days after hospital discharge. Secondary outcomes included a composite measure of in-hospital outcome (death in-hospital or transfer to Uganda's national referral hospital due to progressive severity of illness) and functional status at alive discharge or transfer. Functional status was assessed via the Karnofsky Performance Status (KPS) score.

The primary clinical outcome of the RESERVE-U-2-TOR study was vital status at 60 days after hospital discharge. Secondary outcomes included vital status at 30 days after hospital discharge, functional status at hospital discharge, and functional status at 30 and 60 days after discharge. Functional status was assessed via the KPS score.

For patients discharged or transferred from hospital alive, vital and functional status at the above time points were obtained by contacting patients or their surrogates via telephone.

<u>Clinical Data Collection and Pathogen Diagnostics</u>

At the time of enrollment, all patients in the RESERVE-U-1-EBB and RESERVE-U-2-TOR cohorts underwent clinical assessments and had rapid testing performed for malaria, HIV, and influenza; for persons living with HIV (PLWH) rapid testing for TB was performed. In RESERVE-U-2-TOR, additional testing was performed for whole-blood lactate concentration and SARS-CoV-2 (all patients) and Cryptococcal antigen (PLWH). Testing for these pathogens was informed by World Health Organization (WHO) Integrated Management of Adolescent and Adult Illness guidelines for sepsis and septic shock in resource-limited hospitals in SSA (available at: https://www.who.int/publications-detail-redirect/imai-district-clinician-manual-hospital-care-adolescents-and-adults). These guidelines emphasize rapid testing for malaria and HIV, a low threshold for TB and Cryptococcal testing among PLWH, and consideration of testing or empiric treatment for influenza. SARS-CoV-2 testing was performed given ongoing community transmission during the RESERVE-U-2-TOR study period.

For malaria, rapid testing was performed using qualitative detection of histidine-rich protein II and lactate dehydrogenase of *P. falciparum* in whole-blood samples using the SD Bioline

Malaria AG P.f. and P.f./Pan platforms (Alere/Abbott, Abbott Park, IL, USA). For all patients not known to be living with HIV, HIV testing was performed on whole-blood samples using serial diagnostic platforms (Determine HIV-1/2 Ag/Ab, Alere/Abbott); Chembio HIV 1/2 Stat-Pak, Chembio Diagnostic Systems, Medford, NY, USA), Uni-gold Recombigen HIV-1/2, Trinity Biotech, Ireland). For all enrolled PLWH (known or newly diagnosed), a single urine sample (obtained via urinary catheter or spontaneous void) and a single spontaneously expectorated sputum sample, if obtainable, were tested for evidence of *Mycobacterium tuberculosis* (MTB) infection by the study laboratory technician. For urine samples, 60µL of unconcentrated urine was tested using the Determine TB-LAM Ag assay (Alere/Abbott) as per the manufacturer's recommended operating procedure. The intensity of any visible band on the test strip was graded by comparing it with band intensities on the manufacturer's post-2014 reference card scale; results were considered positive using the grade 1 cutoff. For sputum samples, testing was performed using the Xpert MTB/RIF Ultra platform (Cepheid, Sunnyvale, CA, USA). TB testing for persons without HIV was performed at the discretion of treating clinicians. Nasopharyngeal swab samples were tested at Uganda Virus Research Institute for influenza A and B viruses via real-time polymerase-chain-reaction (RT-PCR).

In RESERVE-U-2-TOR, rapid testing for whole-blood lactate concentration was performed in all patients using the Lactate Plus analyzer (Nova Biomedical, Waltham, MA, USA). Nasopharyngeal swab samples were tested at Uganda Virus Research Institute for SARS-CoV-2 virus via RT-PCR.

For PLWH in RESERVE-U-2-TOR, rapid Cryptococcal antigen testing was performed in serum samples using the CrAg LFA platform (IMMY, Norman, OK, USA). CD4 counts obtained during routine clinical care were recorded. At the Rakai Health Sciences Laboratory in Kalisizo, Uganda, HIV-1 viral load testing was performed on the Abbott RealTime Platform (Abbott Molecular, Inc., Des Plaines, IL, USA). A modified protocol was used to accommodate a lower specimen volume (200ul) with a detection limit of 150 copies. Patients for whom HIV-1 viral loads were undetectable or unquantifiable below this threshold were considered to have viral suppression.

Handling of Missing Clinical Data

As previously described, missing physiologic variables, rapid lactate concentration, CD4 counts, and HIV viral loads (for PLWH) in RESERVE-U-2-TOR were multiply imputed using chained equations (*mice* R package) as per **Tables S1 and S2** (18). Five imputed datasets were reviewed for convergence and plausibility after which one was randomly selected for use. For RESERVE-U-1-EBB, there were no missing physiological variables, rapid lactate and HIV viral load testing were not performed, and routine CD4 testing was rarely performed.

Sample Collection and Storage

At the time of enrollment in both RESERVE-U-1-EBB and RESERVE-U-2-TOR, peripheral blood samples were collected into vacutainer tubes and centrifuged. The resulting serum was aliquoted into cryovials and cryopreserved in liquid nitrogen. Serum samples were then transferred to -80ºC freezers for storage.

Proteomic assays

In cryopreserved serum samples, Olink proteomics (Olink Proteomics AB, Uppsala, Sweden) was performed at the Human Immune Monitoring Center of the Icahn School of Medicine at Mount Sinai (New York, NY, USA) using Target Immunooncology and Cardiometabolic panels. These panels, each of which include 92 proteins, were selected to broadly capture biological domains implicated in the host response to severe infection (i.e., innate and adaptive immune activation and exhaustion, endothelial and cellular metabolic dysfunction, dysregulated coagulation). Comprehensive descriptions of each panel including validation data are available at https://olink.com/products-services/target/immune-response-panel/ and https://olink.com/products-services/target/ cardiometabolic-panel/. For each panel, proteins were quantified via the Olink Proximity Extension Assay. The Proximity Extension Assay uses dual oligonucleotide-labelled antibodies to bind target proteins, creating a unique double stranded DNA barcode which is quantitatively proportional to the initial concentration of the target protein. This barcode then serves as a template for a DNA polymerase-dependent extension step, which is followed by PCR amplification (T100 Thermal Cycle, Bio-Rad Laboratories, Inc., Hercules, CA, USA). Resulting DNA amplicons are quantified by microfluidic qPCR (Fluidigm Biomark HD and JUNO Systems, Standard BioTools Inc., San Francisco, CA, USA) and intensity normalized across batch plates, with relative protein abundance calculated from cycle threshold values and expressed in log2 normalized protein expression (NPX) units by Olink's NPX Manager software. Samples from RESERVE-U-1-EBB and RESERVE-U-2-TOR were analyzed separately; within each cohort, samples were randomized across batch plates and analyzed by technicians blinded to clinical variables and outcomes. Missing NPX values, which were rare, were multiply imputed for each panel using chained equations, predictive mean matching, and all other proteins as predictors (*mice* R package) (18). Five imputed datasets were reviewed for convergence and plausibility after which one was randomly selected for use. Proteins with <20% of NPX values above each panel's estimated limits of detection in either of the RESERVE-U cohorts were excluded from downstream analyses.

**Table S1**: **Missing clinical data for persons living with HIV (PLWH) in RESERVE-U-2-TOR and approach to imputation**

| Variable | Proportion of patients with missing data (%) | Approach to imputation[a] |
|---|---|---|
| WHO HIV clinical stage | 0.0 | N/A |
| CD4 count | 56.6 | MI via chained equations and PMM |
| Use of antiretroviral therapy | 0.0 | N/A |
| HIV-1 viral suppression | 9.0 | MI via chained equations and logistic regression |

Legend: [a]Multiple imputation performed in PLWH using all variables in table as predictors.

Abbreviations: PMM: predictive mean matching, WHO: World Health Organization

**Table S2: Adjusted regression coefficients and 95% confidence intervals (CI) for the association between HIV/TB category and protein expression in the RESERVE-U-1-EBB discovery cohort.** Coefficients and 95% CIs generated from linear regression models including NPX-quantified protein expression as the continuous outcome and HIV/TB phenotype as an ordinal predictor, adjusted for age, sex, illness duration prior to enrollment, and malaria status.

| Protein | Estimate | Statistic | Std Error | p-value | FDR-adjusted p-value |
|---|---|---|---|---|---|
| SERPINA5 | -1.05 | -7.65 | 0.137 | 5.45e -13 | 5.45e -12 |
| IGFBP3 | -0.632 | -6.48 | 0.0976 | 5.64e -10 | 2.82e -9 |
| CRTAM | 0.826 | 6.14 | 0.134 | 3.55e -9 | 1.18e -8 |
| PRSS2 | 1.19 | 5.84 | 0.204 | 1.73e -8 | 4.33e -8 |
| CD70 | 0.732 | 5.78 | 0.127 | 2.41e -8 | 4.83e -8 |
| LAMP3 | 0.834 | 5.11 | 0.163 | 6.70e -7 | 1.12e -6 |
| IL18 | 0.859 | 4.85 | 0.177 | 2.33e -6 | 3.32e -6 |
| EFEMP1 | 0.397 | 4.49 | 0.0883 | 1.12e -5 | 1.40e -5 |
| ICAM3 | 0.356 | 4.08 | 0.0873 | 6.30e -5 | 7.01e -5 |
| IGLC2 | 0.388 | 3.77 | 0.103 | 2.08e -4 | 2.08e -4 |

**Table S3: Adjusted regression coefficients and 95% confidence intervals (CI) for the association between progressive HIV/TB phenotype and protein expression in the RESERVE-U-2-TOR validation cohort.** Coefficients and 95% CIs generated from linear regression models including NPX-quantified protein expression as the continuous outcome and HIV/TB phenotype as an ordinal predictor, adjusted for age, sex, illness duration prior to enrollment, and malaria status.

| Protein | Estimate | Statistic | Std Error | p-value | FDR-adjusted p-value |
|---|---|---|---|---|---|
| IGLC2 | 0.333 | 4.69 | 0.0711 | 4.53e -6 | 4.53e -5 |
| PRSS2 | 0.649 | 3.38 | 0.192 | 8.37e -4 | 4.19e -3 |
| CRTAM | 0.291 | 2.85 | 0.102 | 4.81e -3 | 1.60e -2 |
| SERPINA5 | -0.360 | -2.71 | 0.133 | 7.14e -3 | 1.79e -2 |
| EFEMP1 | 0.230 | 2.42 | 0.0950 | 1.62e -2 | 3.23e -2 |
| CD70 | 0.254 | 2.27 | 0.112 | 2.40e -2 | 4.00e -2 |
| LAMP3 | 0.250 | 2.17 | 0.115 | 3.10e -2 | 4.07e -2 |
| IGFBP3 | -0.234 | -2.15 | 0.109 | 3.25e -2 | 4.07e -2 |
| ICAM3 | 0.117 | 1.35 | 0.0867 | 1.791e -1 | 1.99e -1 |
| IL18 | 0.142 | 1.00 | 0.142 | 3.18e -1 | 3.18e -1 |

**Table S4. Regression coefficients for the association between HIV/TB category and protein expression in PLWH in RESERVE-U-2-TOR cohort.** Each model included NPX-quantified protein expression as the continuous outcome and HIV/TB phenotype as an ordinal predictor. Models were adjusted for CD4 count and HIV-1 viral load suppression.

| Protein | Estimate | Statistic | Std Error | p-value |
|---|---|---|---|---|
| CRTAM | 0.0390 | 0.369 | 0.106 | 0.712 |
| CD70 | 0.0629 | 0.472 | 0.133 | 0.638 |
| IGLC2 | 0.157 | 2.21 | 0.0711 | 0.0289 |
| SERPINA5 | -0.321 | -2.01 | 0.160 | 0.0471 |
| PRSS2 | 0.290 | 1.23 | 0.236 | 0.221 |
| IL18 | -0.0780 | -0.493 | 0.158 | 0.623 |
| ICAM3 | -0.0293 | -0.341 | 0.0861 | 0.734 |
| LAMP3 | 0.0670 | 0.533 | 0.126 | 0.595 |
| EFEMP1 | 0.240 | 2.27 | 0.106 | 0.0250 |
| IGFBP3 | -0.234 | -1.81 | 0.130 | 0.0732 |

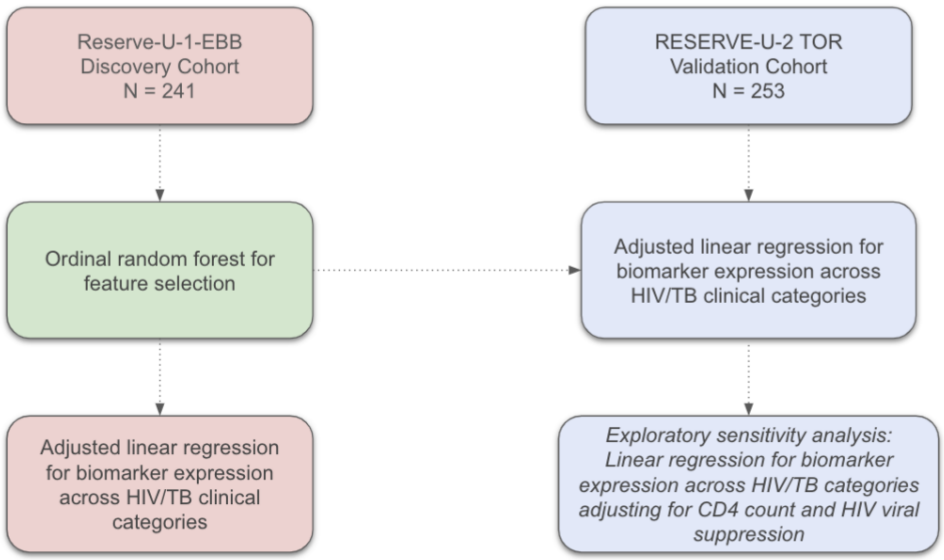


**Supplemental Figure 1.** Overview of study design and analytical workflow for proteomic signature discovery and validation.

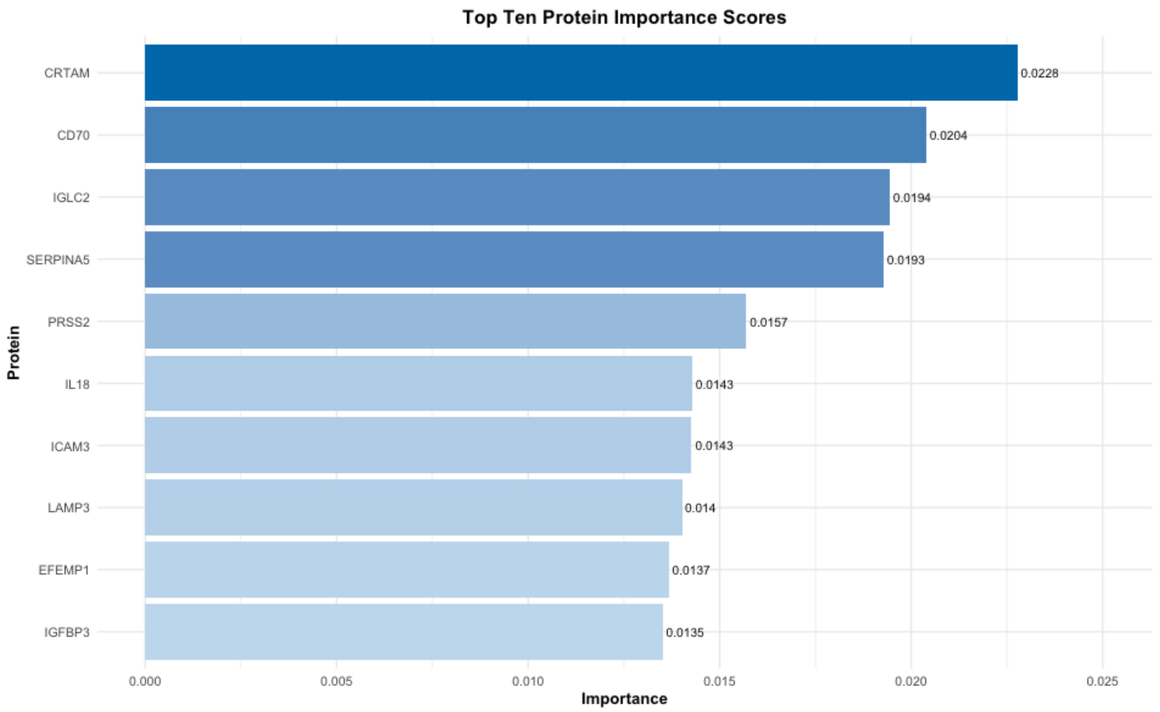


**Supplemental Figure 2.** Bar plot of top 10 proteins most predictive of HIV/TB phenotype in the RESERVE-U-1 EBB discovery cohort. Proteins ranked in descending order of permutation-based variable importance as derived from ordinal random forest models (N = 241).

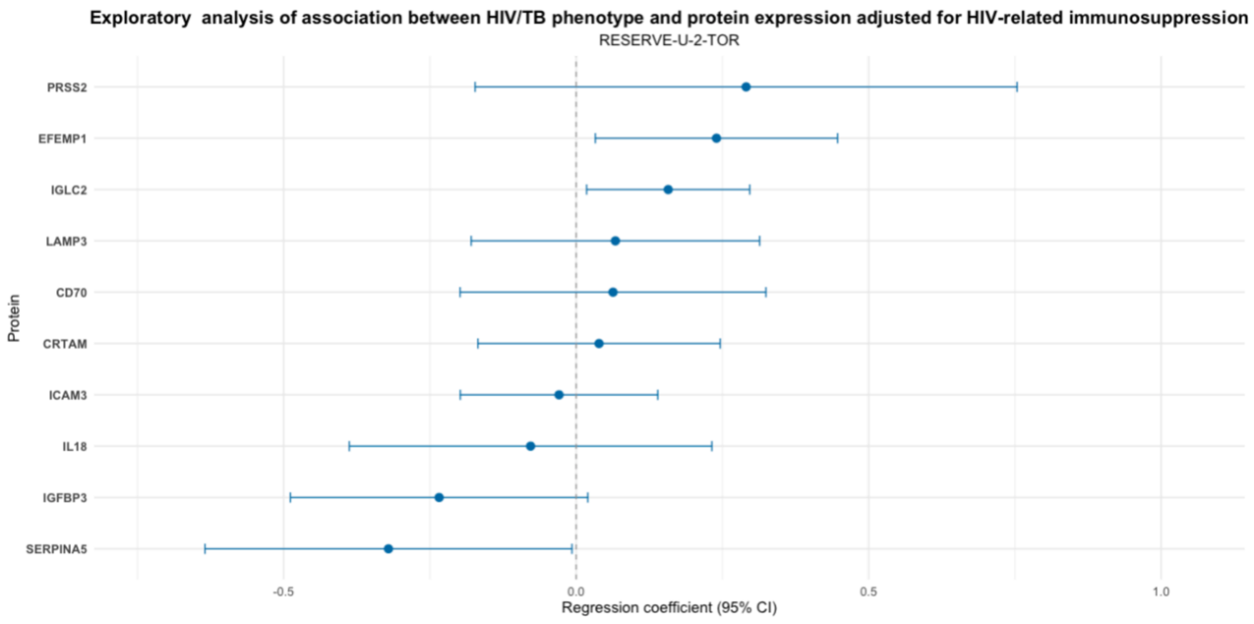


**Supplemental Figure 3.** Forest plot of regression coefficients for the association between HIV/TB phenotype and protein expression in PLWH in RESERVE-U-2-TOR cohort. Each model included NPX-quantified protein expression as the continuous outcome and HIV/TB phenotype as an ordinal predictor. Models were adjusted for CD4 count and HIV-1 viral load suppression (N = 127).
